# Prodrug BMP-7 attenuates pulmonary fibrosis through downregulation of bone marrow derived ApoE+ alveolar macrophage

**DOI:** 10.1101/2024.09.22.24313955

**Authors:** Nam Eun Kim, Sue Bean Cho, Mi Hwa Shin, Hyo Sup Shim, Young Joo Suh, Kim Ha Eun, Jin Gu Lee, Dawool Han, Hyun Kyu Choi, Si Hwan Jang, Sung-Joo Hwang, Nam Hee Kim, Jong In Yook, Hyun Sil Kim, Moo Suk Park

## Abstract

**Background:** Bone morphogenetic protein-7 (BMP-7) antagonises transforming growth factor-β (TGF-β). This study investigated the ability of a prodrug BMP-7, designed as micelle nanoparticles for nasal inhalation, to ameliorate pulmonary fibrosis in a bleomycin (BLM)-induced murine model.

**Materials and Methods:** Fluorescently labelled BMP-7 was delivered to murine lungs via nasal inhalation. Thirty-eight C57BL/6J mice were divided into three groups: control, BLM and BLM with prodrug BMP-7. We then administered the prodrug BMP-7 and vehicle nasally every 72 hours for 21 days. Single-cell RNA sequencing was performed on bronchoalveolar lavage fluid (BALF) from 18 mice, divided into four groups: control, prodrug BMP-7 alone, BLM and BLM with prodrug BMP-7, to assess effects on alveolar macrophages (AM). The expression of ApoE+ AM was compared between normal and idiopathic pulmonary fibrosis (IPF) patients.

**Results:** The prodrug BMP-7 group showed reduced BALF inflammatory cells and significant fibrosis reduction compared to the BLM group. Western blot showed decreased levels of collagen I, α-SMA and fibronectin in the prodrug BMP-7 group, along with downregulation of TGF-β/SMAD signalling. ELISA indicated decreased levels of chemokines CXCL10 and CXCL2 in tissue and BALF. Single-cell RNA sequencing revealed a significant increase in bone marrow-derived ApoE+ AM in the BLM group, which was reduced with prodrug BMP-7. Additionally, ApoE+ expression was higher in IPF patients compared to controls.

**Conclusions:** Prodrug BMP-7 shows potential as a therapeutic agent for pulmonary fibrosis by modulating ApoE+ AM.

## Introduction

Idiopathic pulmonary fibrosis (IPF) is a lethal lung disease characterised by progressive, irreversible fibrosis with a poor prognosis [1]. The excessive accumulation of extracellular matrix and fibrous collagen in the interstitium reduces gas exchange and ultimately causes respiratory distress. Medical treatment has a lower survival rate compared to lung transplantation, there is a growing demand for alternative antifibrotic treatments.

The pathogenesis of IPF is believed to be due to chronic inflammation, but recent findings suggest that the interaction of dysregulated alveolar epithelial cells (AECs) [2–4] with macrophage polarisation [5] and myofibroblast activation are involved. Recent reviews have highlighted that immune cells, especially the macrophages play a crucial role in IPF [5, 6]. Alveolar macrophages (AM) with profibrotic features are analysed through various methods, such as transcriptomic analysis, epigenetics and the association of lipid or senescence metabolism with cellular crosstalk involving nonimmune cells [5, 7]. These macrophages promote fibrosis through the elevated production of TGF-β cytokine [8] and positively regulate lipid metabolism, leading to the generation of foam cells [7]. Single-cell RNA sequencing (scRNA-seq) studies have shown that fibrosis-associated differential gene expression occurs within AM and nonimmune cells (AECs, fibroblasts) [9]. Genes such as Col3a1, Cd302, Mt1, Mt2 and ApoE, known as senescence genes, were remarkably upregulated [10].

Bone morphogenetic proteins (BMPs) play significant roles *via* counter-regulatory mechanisms of TGF-β [11]. TGF-β is recognised as a key fibrogenic cytokine, controlling ECM gene expression through a signalling pathway involving SMAD2/3. It translocate into the nucleus to regulate mesenchymal-type genes [12–14]. In contrast, bone morphogenetic protein-7 (BMP-7) antagonises TGF-β, and previous studies have demonstrated that recombinant BMP-7 (rhBMP-7) can reverse TGF-β-induced pulmonary fibrosis in both *in vivo* and *in vitro* models [15–19].

However, an excessive dose with an initial burst release of rhBMP is required to achieve clinical efficacy in antifibrosis, leading to adverse effects [20]. Animal studies have revealed that current soluble rhBMPs exhibit a half-life of only 7–16 min due to rapid elimination and enzymatic degradation [21]. To overcome the intrinsic limitations of exogenously introduced soluble growth factors, we developed a prodrug BMP-7 that undergoes endogenous protein processing to produce active BMP-7, fused with a protein transduction domain (PTD) [22]. However, PTD-mediated cellular delivery has pharmacological limitations due to the cargo size (<10 KDa) [23].

Therefore, we developed a prodrug BMP-7 in the form of micellised nanoparticles targeting AM, key regulators of alveolar inflammation. Additionally, we investigated its ability to ameliorate pulmonary fibrosis in a preclinical BLM model. The effect of the prodrug BMP-7 on AM was analysed using single-cell transcriptomes of BALF cells. Finally, we investigated ApoE expression, which showed significant changes in scRNA-seq, in IPF human lung tissues compared to normal controls.

## Materials and Methods

### Preparation of prodrug BMP-7

The bacterial expression cassette for the prodrug BMP-7 polypeptide and the purification procedures were conducted as previously described [22]. Briefly, polypeptides were denatured and micellised with filtered 0.1% egg lecithin (BOC Sciences, Shirley, NY, USA) *via* sonication. The typical micelle size, as determined by direct light scattering, was approximately 180–200 nm. The endocytic transduction and exosomal secretion of active BMP-7 have been well established in a previous study [24, 25].

### In vivo distribution analysis

C57BL/6 mice were intranasally instilled with indocyanine green (ICG)-labelled micellised prodrug BMP-7 (10 μg) and subjected to in vivo fluorescent imaging at different time points. Fluorescent images were obtained using VISQUE® InVivo Smart (Vieworks, Anyang, South Korea) and measured at 30 min, 1 h, 6 h, 24 h, 48 h and up to 72 h. In the ex vivo experiment, mice were sacrificed on day 7 after inhalation of prodrug BMP-7 (1 μg), and ICG-labelled prodrug BMP-7 in the lung tissue was analysed. All imaging data were processed using CleVue™ software (Vieworks).

### Experimental groups

Eight-week-old male C57BL/6 mice weighing 22–25 g were purchased from Orient Bio (Sungnam, Republic of Korea). Mice were maintained on a 12-hour light/dark cycle with access to food and water. Thirty-eight mice were divided into three groups: (A) phosphate-buffered saline (PBS) inhalation and vehicle group (control group) (*n* = 12), (B) BLM inhalation and vehicle group (BLM group) (*n* = 13), and (C) BLM-induced pulmonary fibrosis group with prodrug BMP-7 inhalation (BLM + prodrug BMP-7 group) (*n* = 13).

In the control group, 50 μl of sterilised PBS was instilled intranasally. In the other groups, BLM (Sigma, St. Louis, MO, USA) at 3 U/kg dissolved in 50 μl of PBS was administered in the same manner. Mice were anesthetized using isoflurane inhalation (Abbott Laboratories) in a supine position, and the solution was delivered *via* microsyringe (Hamilton Company, Cat. number 7637-01) with intranasal instillation. The release rate was adjusted so that the mice could inhale the solution without forming bubbles, similar to the method proposed by Leem et al [26]. As a posttreatment, prodrug BMP-7 (1.0 μg/g dissolved in 50 μl) or vehicle was inhaled intranasally three times weekly for 3 weeks.

### Isolation of bronchoalveolar lavage cells and cell counts

All mice were euthanised 21 days after BLM/PBS injection, and bronchoalveolar lavage fluid (BALF) was collected through a tracheal cannula with two 1 ml aliquots of sterile saline. Samples were centrifuged for 10 min at 1,500–5,000 rpm at 4°C. The cell pellet was then resuspended in 100 μL of PBS, and cell counting was quantitatively and qualitatively analysed using a haemocytometer (Marienfeld, Germany). The slides were retrieved, desiccated and stained by immersion in Diff-Quik (Sysmex Corporation). The protein content of the BALF supernatant was estimated using Coomassie Brilliant Blue G-250 (Quick Start^TM^ Bradford Protein Assay), and the absorbance was read at 562 nm using a spectrophotometer (Supplementary Annex).

### Lung tissue harvest and histologic examination

Before protein extraction, the right lung was stored at −80°C after flushing the pulmonary vasculature with saline. The left lung was inflated with low-melting-point agarose (4%) in PBS at 25 cm H_2_O pressure until the edges of the pleura became sharp. The tissues were then sliced, fixed for 24 h in 10% formaldehyde in PBS, embedded in paraffin and sectioned at 5 μm. Subsequently, slides were stained with hematoxylin and eosin (H&E) and Masson’s trichrome. Moreover, two qualified investigators evaluated pulmonary fibrosis semi-quantitatively using the modified Ashcroft score scale, which ranges from 0 to 8 [27]. Pulmonary fibrosis was recorded as dominant in any area where it occupied more than 50% of the fields. The final score was presented as the mean of the scores observed in all fields.

### Western Blotting

The frozen lungs were automatically homogenised in 600 μL of homogenisation buffer (PROPREP^TM^ Extraction Solution), and cell lysis was induced by incubation for 20 min at −20°C. The samples were then centrifuged at 4°C, 13,000 × g for 30 min. The antibodies used in this study included α-SMA (Abcam, Cat. No. ab7817, 1:2,000), collagen I (Abcam, Cat. No. ab34710, 1:2,000), fibronectin (Abcam, Cat. No. ab2413, 1:2,000), β-actin (Santa Cruz, Cat. No. sc-47778, 1:1,000), p-Smad2/3 (Cell Signalling, Cat. No. 8828, 1:500), and Smad2/3 (Cell Signalling, Cat. No. 3102, 1:500) (Supplementary Annex).

### Enzyme linked immunosorbent assay

The levels of C-X-C motif chemokine 10 (CXCL10), C-X-C motif chemokine 2/matrix metalloproteinase-2 (CXCL2/MMP-2), interleukin-1 β (IL-1β) and interleukin-6 (IL-6) in whole-lung homogenates and BAL fluid were measured using quantified ELISA kits (Cat. No. RND-LXSAMSM-05_Mouse Premixed Multi-Analyte Kit) according to the manufacturer’s instructions. The cytokines used in this study included CXCL10, CXCL2/MMP-2, IL-1β and IL-6 (R&D Systems).

### Single-cell RNA-seq analysis of BALF cells of mouse & human IPF blood samples

BALF scRNA-seq analysis was conducted in four groups. Eighteen mice were divided into four groups, with the addition of the prodrug BMP-7-alone group: PBS group (control group) (*n* = 6) (G1), prodrug BMP-7 group (*n* = 6) (G2), BLM group (*n* = 3) (G3) and BLM + prodrug BMP-7group (*n* = 3) (G4).

BALF single cells were collected and pooled for each group. In total, 40,000 cells per group were loaded onto microwell cartridges of the BD Rhapsody Express system (BD). Single-cell whole transcriptome analysis libraries were prepared using the BD Rhapsody WTA Reagent Kit (BD: 633802) according to the manufacturer’s instructions. The final indexed PCR libraries were sequenced on Illumina HiSeq using the High Output Kit v2.5 (150 cycles, Illumina) for 2 × 75 bp paired-end reads with an 8 bp single index.

The FASTQ-format sequencing raw data were processed through the BD Rhapsody WTA Analysis pipeline (version 1.0, Revision 6) on the SevenBridges Genomics online platform (SevenBridges). The resulting expression matrix was used for further data analysis. Unless otherwise specified, data normalisation, dimensionality reduction and visualisation were performed using the Seurat package (version 4.3.0). Cells were filtered based on the following criteria to ensure data quality: having 500– 6,000 genes per cell (nFeature_RNA) and a percentage of mitochondrial genes (percent.mito) of <25%. For the unsupervised hierarchical clustering analysis, the FindClusters function in the Seurat package was implemented. Various resolutions of 0.1–0.9 were tested, and the final resolution was selected based on the most stable and relevant outcome, utilising the clustree R package and considering prior knowledge (Supplementary annex).

### Immunofluorescence & Immunohistochemistry staining

ApoE recombinant rabbit monoclonal antibody (Thermo Fisher Scientific, Catalogue No. 701241) and the macrophage marker, CD68 (Santa Cruz Biotechnology, Inc., Catalogue No. sc-20060), were diluted in DAKO antibody dilution buffer (1:1,000) and incubated overnight at 4°C. Goat antirabbit IgG secondary antibody, Alexa Fluor^TM^ 488, or goat antimouse IgG secondary antibody, Alexa Fluor^TM^ 568 (Invitrogen), was diluted 1:1,000 and incubated for 1 h at room temperature. After washing the secondary antibodies with PBS, nuclei were stained for 20 min with 1 μg/ml DAPI.

Fluorescence images were obtained using a Zeiss LSM700 confocal microscope (Carl Zeiss, Berlin, Germany). Morphological analysis of the confocal images was performed using Metamorph microscopy analysis software (version 7.1, Molecular Devices, Sunnyvale, CA, USA).

## Statistics

Data were compared an using unpaired Student’s *t*-test or ANOVA with Bonferroni’s multiple comparisons in Prism 5.0 (GraphPad Software, Durham, NC, USA). Data were reported as mean ± SD, and differences were considered significant at *P* < 0.05.

## Study approval

Animal protocols were approved by the Institutional Animal Care Committee, Yonsei University College of Medicine (2021-31-0820) and followed the recommendations of the Guide for the Care and Use of Laboratory Animals by the National Institutes of Health.The IPF patient registry comprised paraffin-embedded lung tissue samples collected from patients who underwent lung transplantation due to IPF. Case 1 was a IPF patient who underwent left lung transplantation in August 2022. Case 2 was a IPF patient who underwent bilateral lung transplantation in August 2022. The study protocols were approved by the Institutional Review Board of Severance Hospital (Approval no. 4-2013-0770).The normal control group included patients who underwent thoracic surgery due to pneumothorax, and normal lung tissue samples were obtained. The study protocols were approved by the Institutional Review Board of Severance Hospital (Approval no. 4-2019-0447).

Bulk transcriptomics was performed and analyzed on the blood of 5 normal healthy patients and 20 IPF patients. The study protocols were approved by the Institutional Review Board of Severance Hospital (Approval no.: 4-2020-0229).This study adhered to the principles of the Declaration of Helsinki (2000) and the Declaration of Istanbul (2008).

## Results

### Prodrug BMP-7 is delivered to the alveoli and shows prolonged tissue retention

We performed a lung distribution dosing study in an *in vivo* and *ex vivo* mouse model to determine whether the inhaled prodrug BMP-7 is delivered to the small airways and alveoli and retained in pulmonary tissue. Nine C57BL/6 mice were intranasally instilled with ICG-labelled prodrug BMP-7 (1 and 10 μg) and imaged using the IVIS system at different time points: 30 min, 1 h, 6 h, 24 h, 48 h and up to 72 h *in vivo*. The inhaled prodrug BMP-7 was retained in the lung tissue for up to 72 h at a dose of 10 μg (Figure 1A).

**Figure 1.**
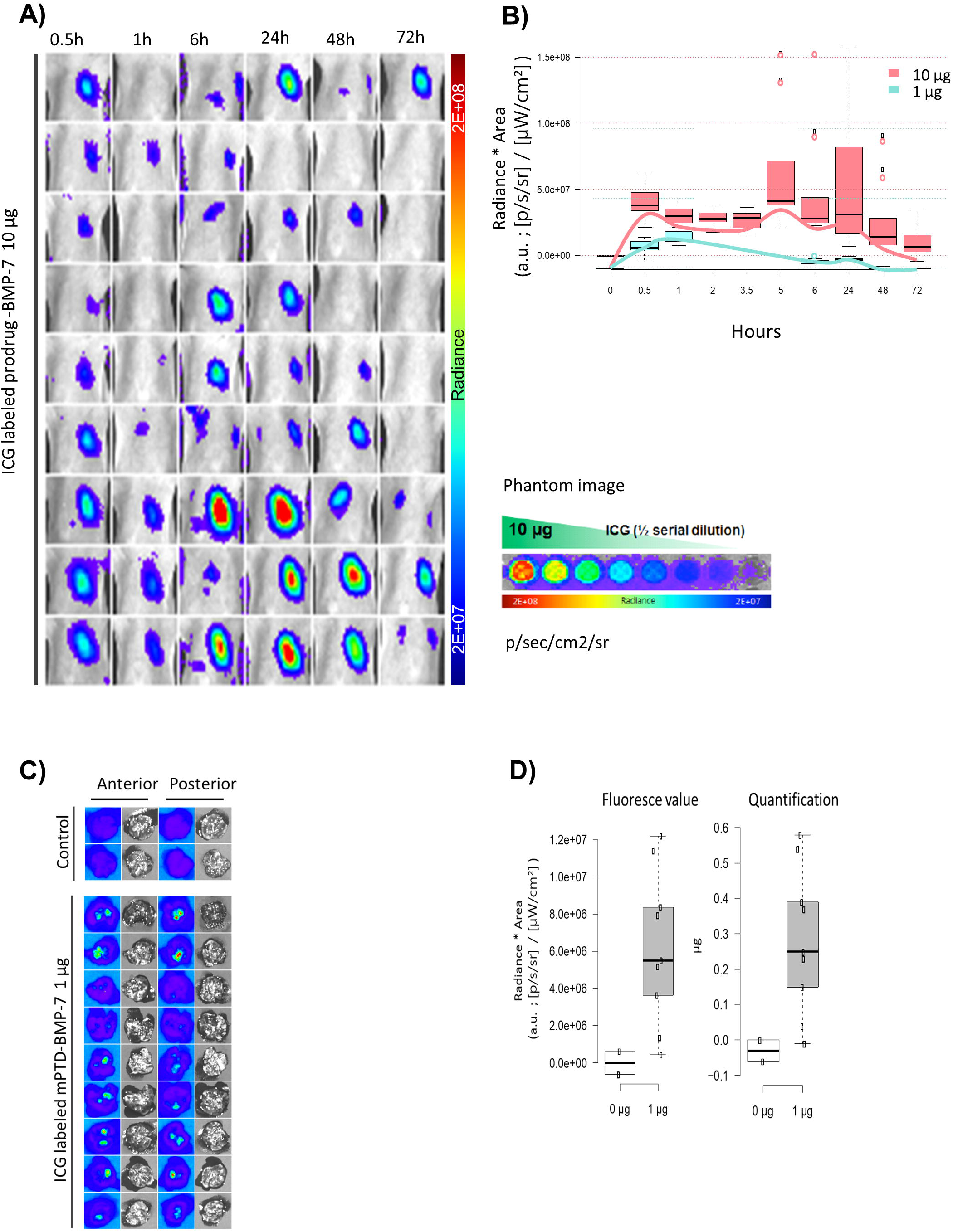
*In vivo* and *Ex vivo* IVIS imaging. (A) *In vivo* IVIS images of C57BL/6 mice with intranasal instillation of prodrug BMP-7 (10 μg). (B) Inhalation of different concentrations of prodrug BMP-7 with 1 and 10 μg, respectively, within 72 h. (C) *Ex vivo* experiment with ICG-labeled prodrug BMP-7 (1 μg) after 7 days of inhalation exposure. (D) Comparison of fluorescence values and radiation quantification between the control and experimental groups (prodrug BMP-7, 1 μg inhalation)

Figure 1B shows the results of experiments with different concentrations (1 and 10 μg). The drug reached its maximum level within the first hour after inhalation and remained in the lung tissue for approximately 48 h in the 1 μg group (*n* = 9) and 72 h in the 10 μg group (*n* = 9). Higher drug concentration resulted in higher radiation intensity and longer duration. Furthermore, in the *ex vivo* prodrug BMP-7 distribution analysis, an autopsy was performed on day 7 after inhalation of 1 μg of prodrug BMP-7. The remaining drug in the lung tissue was analysed using IVIS. Prodrug BMP-7 remained in the lung tissue even after 1 week of nasal inhalation compared with the control group (Figure 1C and D).

In addition to the *in vivo* and *ex vivo* distribution analysis results, the effective drug concentration and the number of inhalations were determined to be 1 μg and repeated instillation within 72 h, respectively. Moreover, immunofluorescence staining was performed 24 h later in the mouse group that inhaled the nanoparticle prodrug BMP-7, confirming that the prodrug BMP-7 was delivered to the alveoli and small airways *via* inhalation (Supplementary Figure 1).

### Prodrug BMP-7 attenuates BLM-induced pulmonary fibrosis

Figures 2A and B show that BLM-induced inflammatory cell infiltration, increasing the total BALF cell count. In contrast, inhaled prodrug BMP-7 decreased BALF inflammatory cells and total cell counts (29.25 × 10^4^ in the control group, 50.4 × 10^4^ in the BLM group, and 10.5 × 10^4^ in the prodrug BMP-7 group; *P* < 0.001). Additionally, Figure 2C shows histological changes related to pulmonary fibrosis observed in H&E and Masson’s trichrome staining. Lung tissue exposed to PBS was well-organised and free of inflammatory cells and collagen deposits in the alveoli. BLM-exposed tissues showed disordered structures, thickened interalveolar septa, inflammatory cell infiltration and collagen deposits in the interstitium. Conversely, inhaled prodrug BMP-7 decreased collagen matrix density, as shown by Masson’s trichrome staining and quantitatively measurement of total collagen content using ImageJ. Moreover, the modified Ashcroft lung fibrosis score was increased in the BLM group compared with the control group, and prodrug BMP-7 inhalation improved the fibrosis score (0.00 in the control group, 5.00 in the BLM group, and 3.00 in the inhaled prodrug BMP-7 group; *P* < 0.001) (Figure 2D).

**Figure 2.**
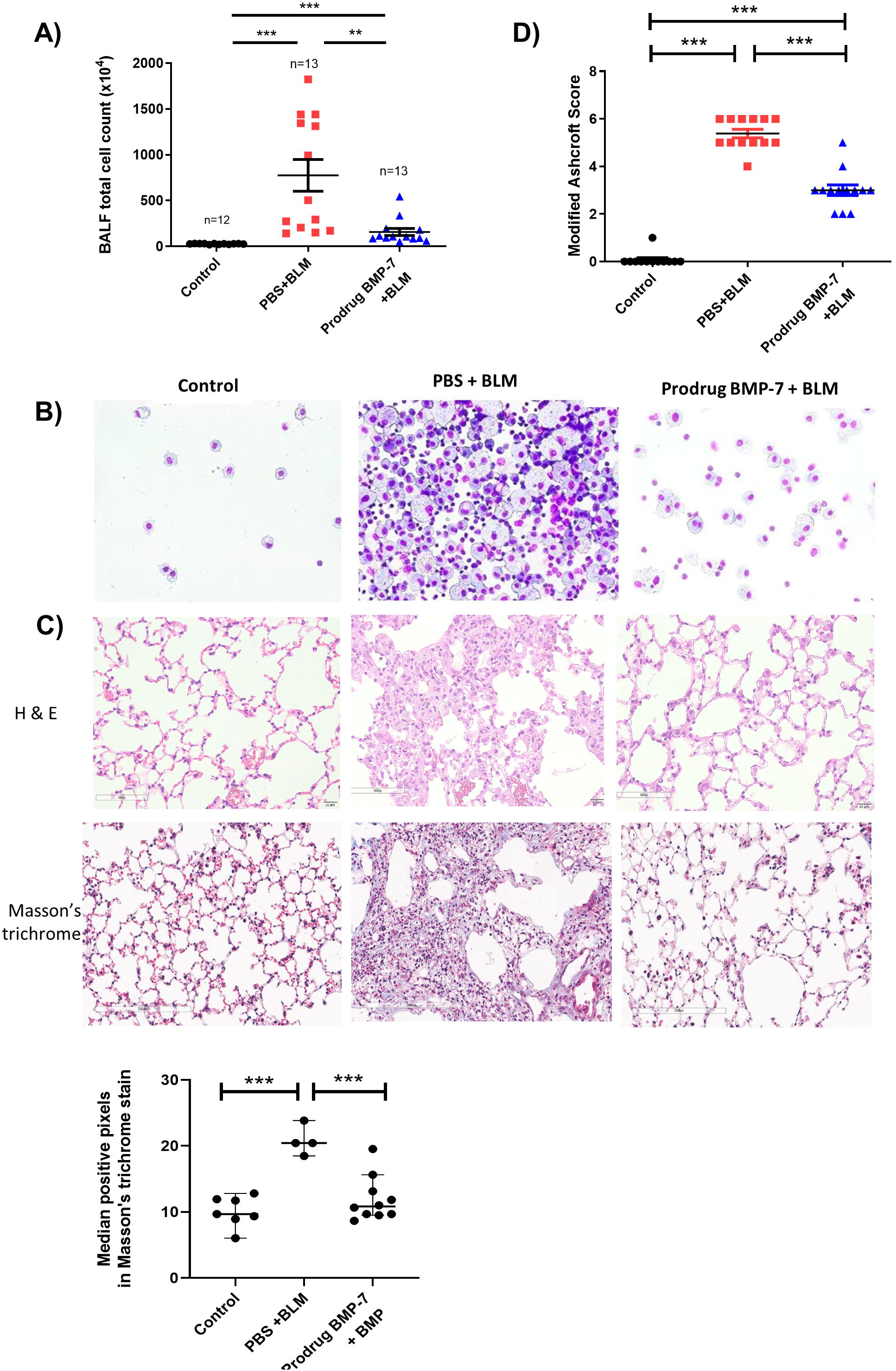
Inhaled prodrug BMP-7 attenuates BLM-induced lung fibrosis. (A) Cell counts and (B) cytology in BALF (C) Lung tissues stained with H&E and Masson’s trichrome stain, magnification 200x, total collagen count measured by ImageJ pixels. (D) Modified Ashcroft lung fibrosis scores in mice lung tissue. Micellized prodrug BMP-7 decreased inflammatory cells, fibrotic lesions, and modified Ashcroft lung fibrosis score compared with those in the BLM group. * *P* < 0.05; ** *P* < 0.01; and *** *P* < 0.001.

### Prodrug BMP –7 inhibits the TGF-**β**/Smad2/3 signalling pathway in BLM-induced pulmonary fibrosis

The antifibrotic effect of prodrug BMP-7 was characterised by a lower level of ECM proteins, such as collagen I, fibronectin and α-SMA, compared with the BLM group (*P* < 0.05; Figure 3A–D). Given the antifibrotic effect of prodrug BMP-7, we investigated whether the mechanism involves the downregulation of TGF-β/Smad 2/3 signalling. Therefore, the expression of TGF-β/Smad 2/3 signalling was measured in all groups. As measured by Western blotting on day 21, BLM stimulated the expression of phosphorylated Smad 2/3 relative to the control group. Prodrug BMP-7 inhalation attenuated the expression of phosphorylated Smad 2/3 (*P* < 0.001; Figure 3E). These results revealed that prodrug BMP-7 attenuated pulmonary fibrosis by downregulating TGF-β/Smad 2/3 signalling.

**Figure 3.**
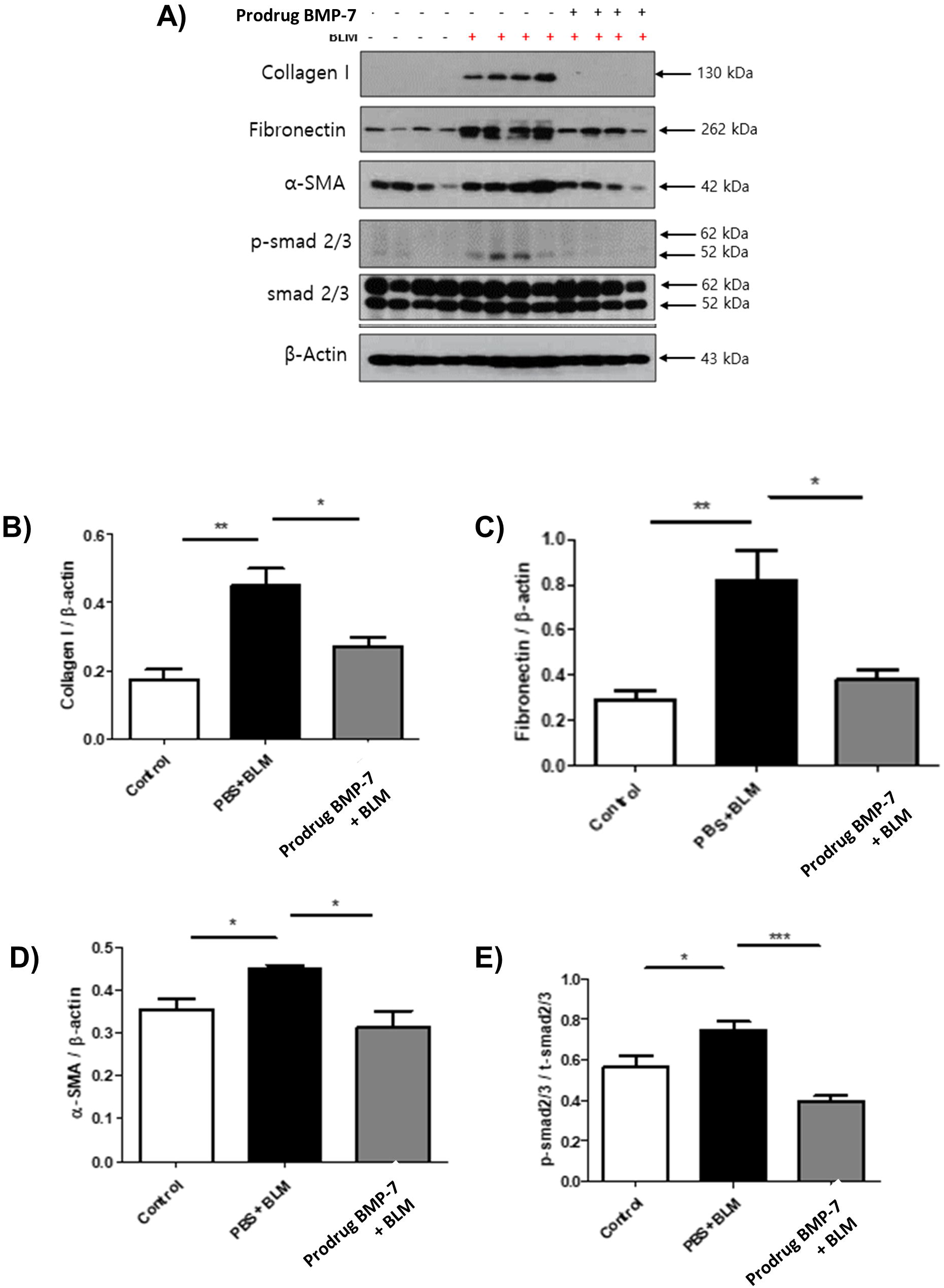
Prodrug BMP-7 attenuates BLM-induced pulmonary fibrosis by downregulating TGF-β/Smad 2/3 pathway. Fibrosis markers and Smad protein were detected using Western blotting in mouse lung tissue to assess the antifibrosis effect of prodrug BMP-7 (Figure 3A). Prodrug BMP-7 suppressed the expression of collagen I (3B), fibronectin (3C), α-SMA (3D), and p-Smad 2/3 (3E). The data represent the means ± SD. * *P* < 0.05; ** *P* < 0.01; and *** *P* < 0.001.

### Prodrug BMP-7 attenuates BLM-induced pulmonary fibrosis by suppressing CXCL10, CXCL2/MIP-2, IL-1**β** and IL-6

To determine the antifibrotic effect of prodrug BMP-7, we evaluated the levels of CXCL10, CXCL2/MIP-2, IL-1β and IL-6 using ELISA on day 21 with mouse lung tissue lysates. CXCL10, CXCL2/MIP-2, IL-1<u>β</u> and IL-6 levels significantly increased after exposure to BLM. However, these levels decreased after inhalation of prodrug BMP-7 (Figure 4A–D). The same result was observed in BALF (Figure 4E–F).

**Figure 4.**
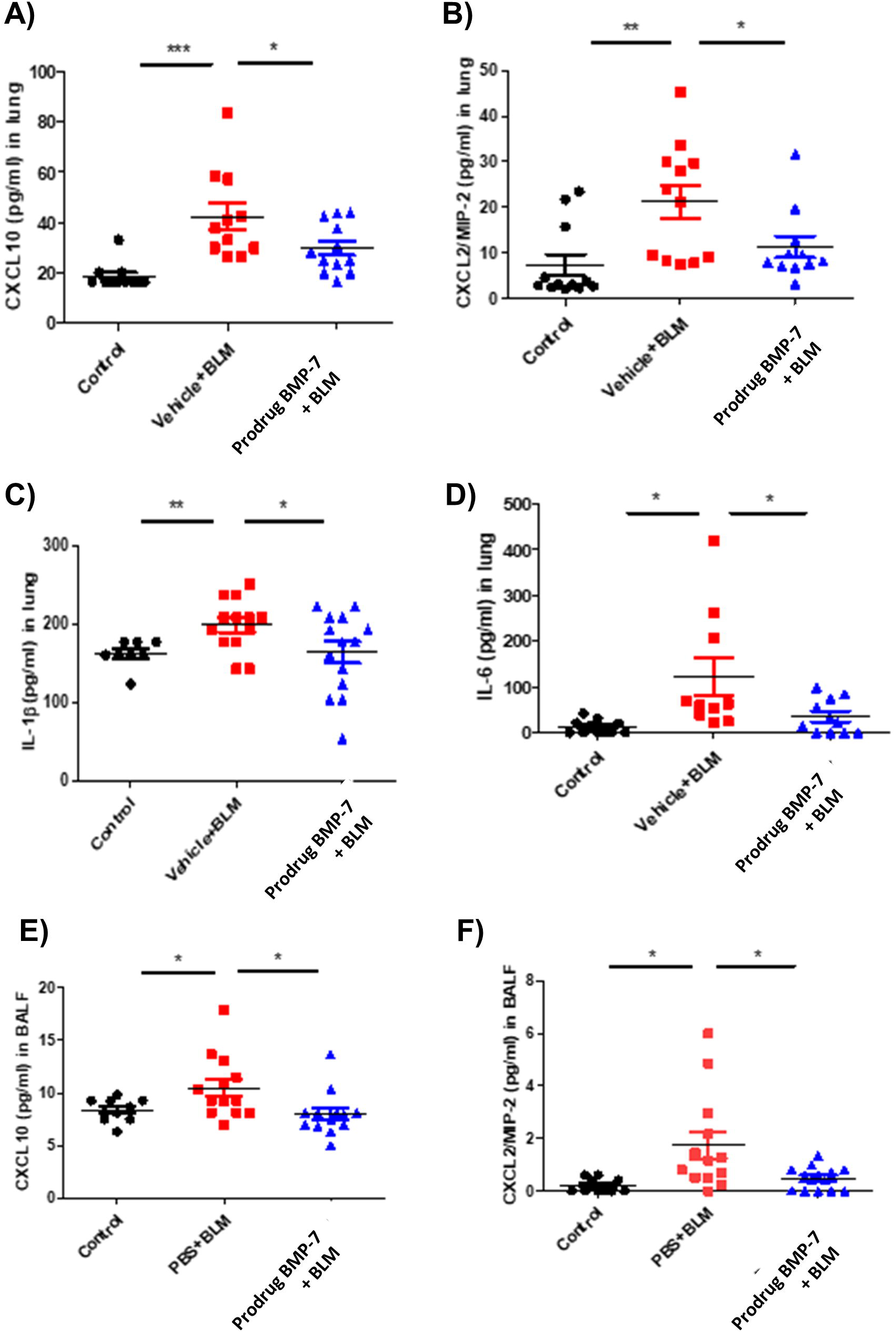
CXCL10, CXCL2/MMP-2, IL-1β, and IL-6 levels in lung tissue lysates and BALF fluid. Cytokine levels measured using ELISA decreased in mice with prodrug BMP-7 inhalation compared with those in the BLM-induced lung injury group. CXCL10, CXCL2/MMP-2, IL-1β, and IL-6 (A–D) in lung tissue lysates (E–F) of BALF. Values are presented as means ± SD * *P* < 0.05; ** *P* < 0.01; and *** *P* < 0.001.

### Prodrug BMP-7 changes the composition of inflammatory cells in the BALF of the BLM-induced lung fibrosis mouse model

To analyse the effect of prodrug BMP-7 on single-cell transcriptomes of BAL fluid cells in vivo, we designed a mouse lung fibrosis model induced by BLM inhalation, followed by nasal inhalation of prodrug BMP-7. The mice were divided into four groups: the PBS group (control group) (G1), the prodrug BMP-7 group (G2), the BLM group (G3) and the BLM + prodrug BMP-7 group (G4) (Figure 5A). scRNA-seq was performed using the BD Rhapsody platform. After removing doublets, 97,219 filtered cells were analysed using the uniform manifold approximation and projection (UMAP) algorithm. Each cluster was identified using representative marker genes for each cell type (Figure 5B and Supplementary Figure 2). The proportion of each cell type demonstrated differences between groups (Figure 5C). Notably, the proportion of macrophages decreased in the BLM treatment group (G3) compared with the control groups (G1 and G2). However, it exhibited an increase in the BLM+ prodrug BMP-7 treatment group (G4) compared with G3, while the proportion of T/NK cells changed reciprocally.

**Figure 5.**
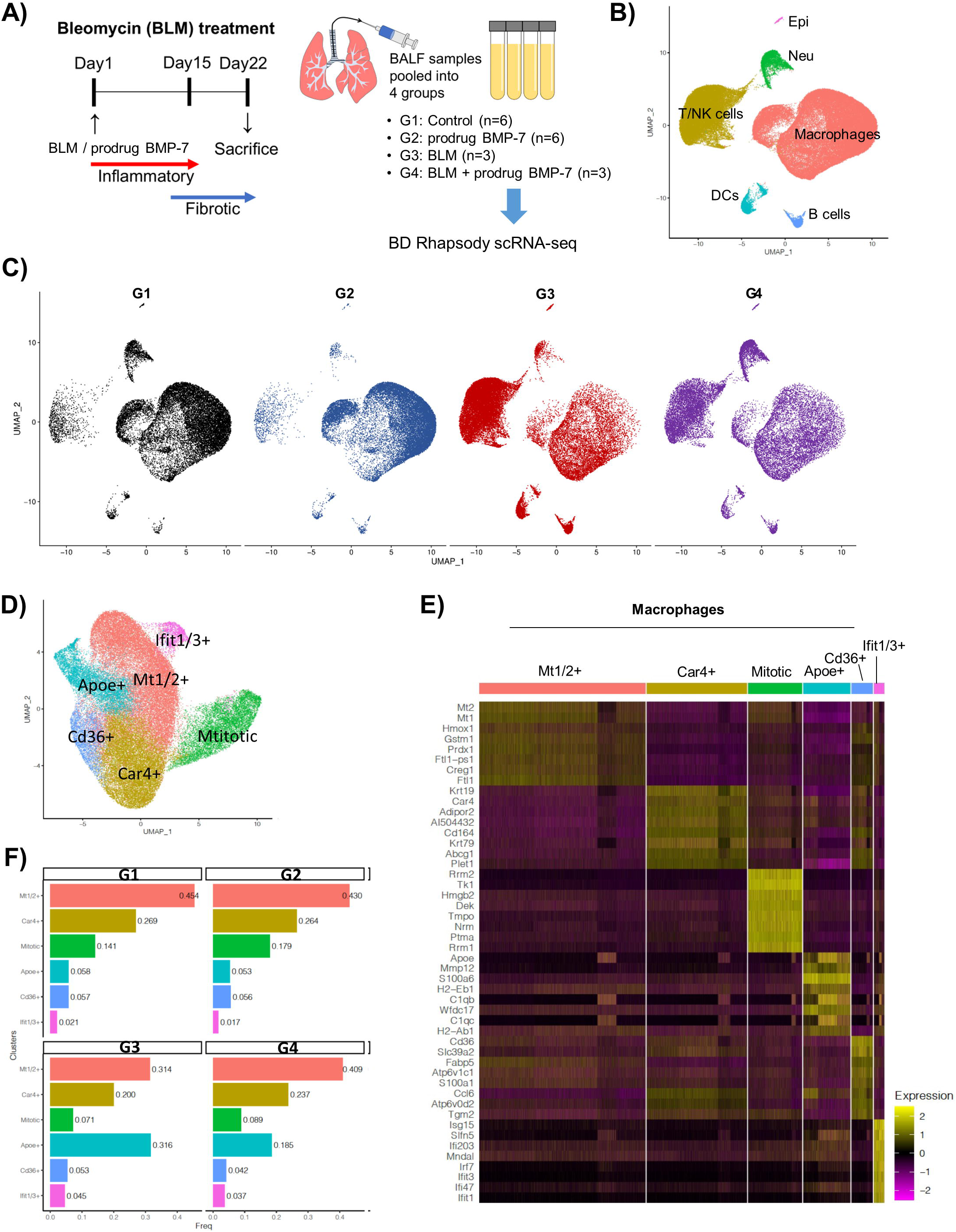
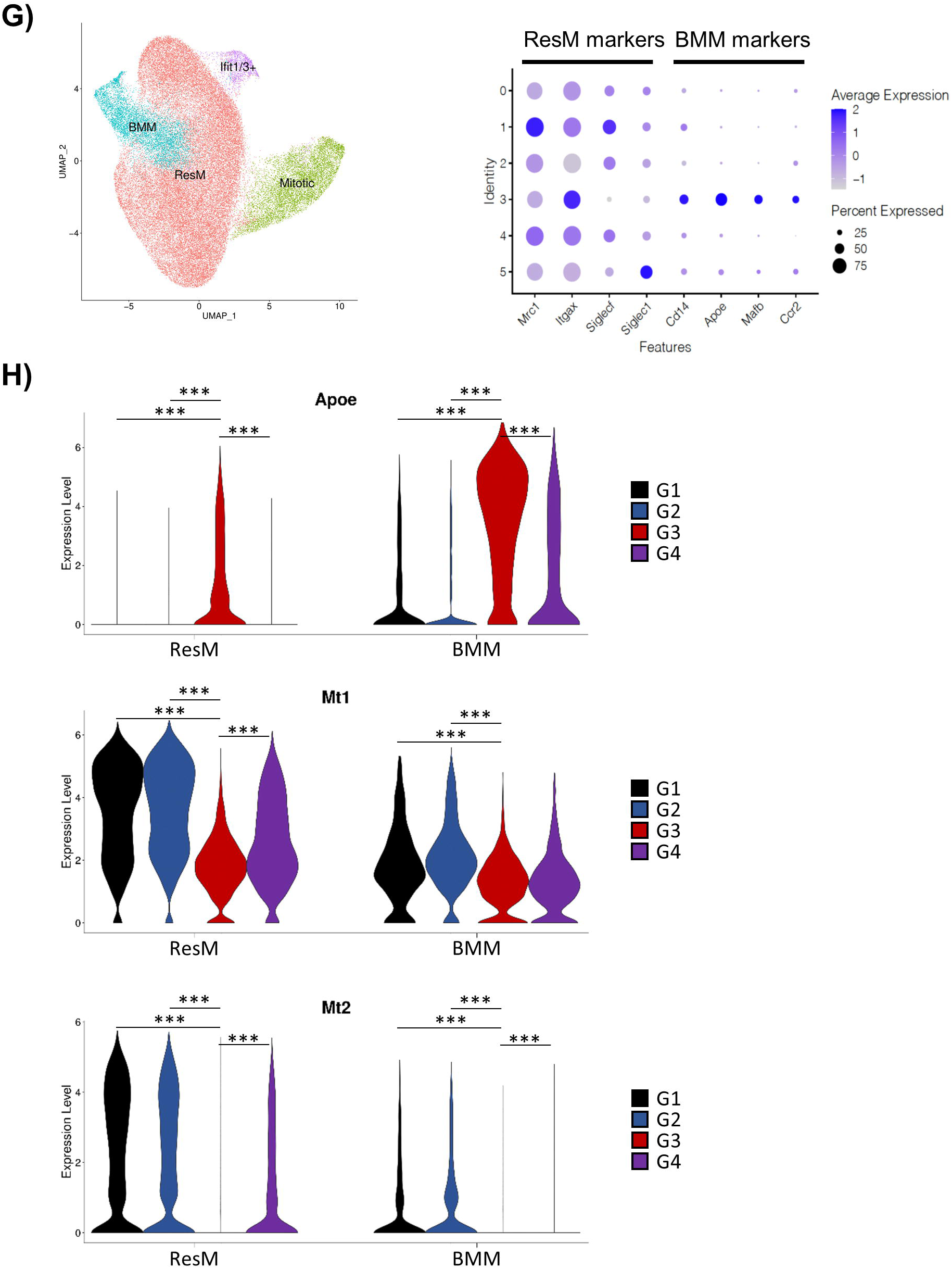
Prodrug BMP-7 decrease inflammatory cells in the BALF of BLM induced lung fibrosis mouse model. (A) Schematic of experimental design for single-cell transcriptomes of BALF cells. (B) Cell-type annotations projected into UMAP space. (C) UMAP colorized for and separated by control (black), prodrug BMP-7 (dark blue), BLM (red), and BLM with prodrug BMP-7 (purple) of BALF scRNA-seq data. (D) UMAP visualization of macrophage subclusters. (E) Heatmap of top marker genes for macrophages with normalized expression colorized (low expression, purple; high expression, yellow). (F) Bar plots of the proportion of macrophage subclusters in each group. (G) UMAP plot of reclassified macrophage subclusters using ResM and BMM markers. (H) Violin plots of ApoE, Mt1, and Mt2 gene expression in ResM and BMM across each group.

Conversely, no significant difference was observed between the prodrug BMP-7 group (G2) and the control group (G1). Moreover, cells designated as macrophages were isolated for subpopulation analysis to explore the role of macrophages in BLM-induced lung fibrosis (Figure 5D). A total of 57,293 macrophages were divided into six subclusters, each subcluster named using one of the top eight differentially expressed genes (DEGs) (Figure 5E): Mt1/2+ macrophages, Car4+ macrophages, mitotic macrophages, ApoE+ macrophages, Cd36+ macrophages and Ifit1/3+ macrophages. Compared with the control groups (G1 and G2), the proportion of ApoE+ macrophages significantly increased in the BLM treatment group (G3) and decreased in the prodrug BMP-7 and BLM treatment group (G4) (Figure 5F). A volcano plot of DEGs revealed a mirror-like pattern between G3 *versus* G1 and G4 *versus* G3 (Supplementary Figure 3). Fewer DEGs were observed in G2 *versus* G1, G4 *versus* G1 and G4 *versus* G2. Violin plots showed differential gene expression among G1, G3 and G4 (Supplementary Figure 4). In each subcluster, ApoE, C1qb and H2-Eb1 increased in G3 compared with the control groups (G1 and G2) and then decreased in G4. Conversely, Mt1 and Mt2 decreased in G3 and increased in G4.

ApoE+ macrophages, exhibiting high levels of Cd14, ApoE, Mafb and Ccr2, were designated as bone marrow-derived macrophages (BMM). The remaining macrophage subtypes, excluding mitotic and Ifit1/3+ macrophages (Mt1/2+ macrophages, Car4+ macrophages and Cd36+ macrophages) were combined as resident macrophages (ResM). Trends of increase and decrease in DEGs are presented in Figure 5G. ApoE and C1qb in ResM and BMM increased in G3 compared with the control groups (G1 and G2) and then decreased in G4. H2-Eb1 in ResM followed a similar pattern to ApoE and C1qb, while in BMM, it increased in G3 and decreased in G4. Mt1 in ResM and Mt2 in ResM and BMM decreased in G3 and increased in G4, whereas Mt1 in BMM showed little change in G4 (Figure 5H and Supplementary Figure 5).

Gene set enrichment analysis identified the top 10 and bottom 10 enriched pathways using DEGs between G1 and G3 and G3 and G4 in ResM and BMM, based on the normalised enrichment score values (Supplementary Figures 6 and 7). In G3, genes related to coagulation and complement pathways increased, while they decreased in G4.

### Prodrug BMP-7 downregulated ApoE expression with macrophages in a BLM-induced fibrosis mouse model and normal and IPF human lung tissues

Immunofluorescence and immunohistochemistry staining were performed to evaluate the role of prodrug BMP-7 in ApoE expression on AM. Figure 6A shows that macrophages were exclusively expressed in mice treated with the prodrug BMP-7 vehicle. The BLM group exhibited increased ApoE expression within the cytoplasm of AM (Figure 6B), while the prodrug BMP-7 group showed a marked decrease in ApoE expression on AM (Figure 6C), indicating possible downregulation of ApoE expression in response to prodrug BMP-7. Furthermore, the response of ApoE expression on AM in normal and IPF human lung tissues was investigated. Figures 7A and B show that in the patient’s normal lung tissue, ApoE immunostaining indicated the absence of cytoplasmic ApoE expression. Additionally, the bronchioles showed exclusive expression of macrophages with no detectable cytoplasmic ApoE expression (Figures 7C and D). However, in the IPF lung tissue, ApoE immunostaining showed robust and active ApoE expression, observed in the bronchioles, alveolar sacs and within the interstitium (Figures 7E–J). The IPF patient in case 1 (Figures 7E and F), histology exhibited marked fibrosis with fibroblastic foci, airway dilatation and moderate inflammation, consistent with the pathological findings of usual interstitial pneumonia. The IPF patient in case 2 (Figures 7I and J), pathology revealed marked fibrosis and moderate inflammation with emphysema-like bullae and fibroblastic foci. Immunofluorescence staining was performed to elucidate the functional role of ApoE and explore the intricate interplay of cells and proteins in the gas exchange region, particularly its involvement in inflammatory and immune responses with macrophages (Figure 7G, H, K and L). The analysis revealed strong ApoE expression in the cytoplasm surrounding the nucleus, co-localising with the presence of macrophages, indicating that ApoE expression is induced and promoted in human pulmonary fibrosis.

**Figure 6.**
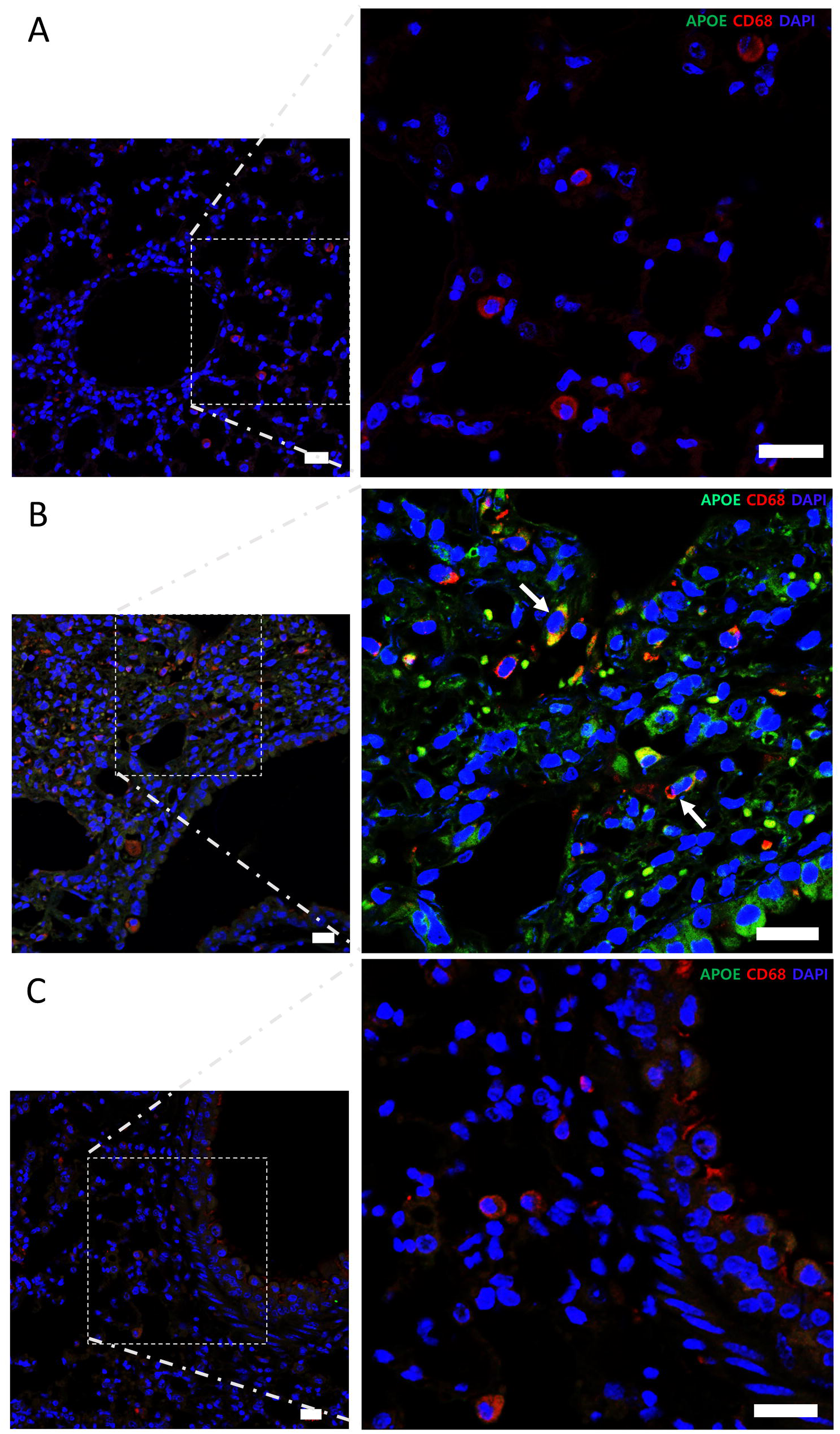
A group of mice with BLM-induced fibrosis was further treated with prodrug BMP-7 to determine its effect on ApoE expression. (A) In the group of mice treated with the vehicle of the prodrug BMP-7, exclusive expression of macrophages was observed (shown in red). (B) In the group of mice treated with BLM alone, cells co-expressing ApoE with macrophages exhibited cytoplasmic localization (highlighted by white arrows), accompanied by a significant increase in ApoE expression within the cytoplasm (shown in green). (C) In a group of mice treated with prodrug BMP-7 in combination with BLM, a distinct pattern was observed. ApoE expression was markedly decreased in the treated group, except for certain cells, particularly macrophages (shown by red fluorescence), indicating a possible downregulation of ApoE expression in response to prodrug BMP-7 in the presence of BLM-induced fibrosis. Cell nuclei were counterstained with DAPI (blue); scale bar: 20 μm

**Figure 7.**
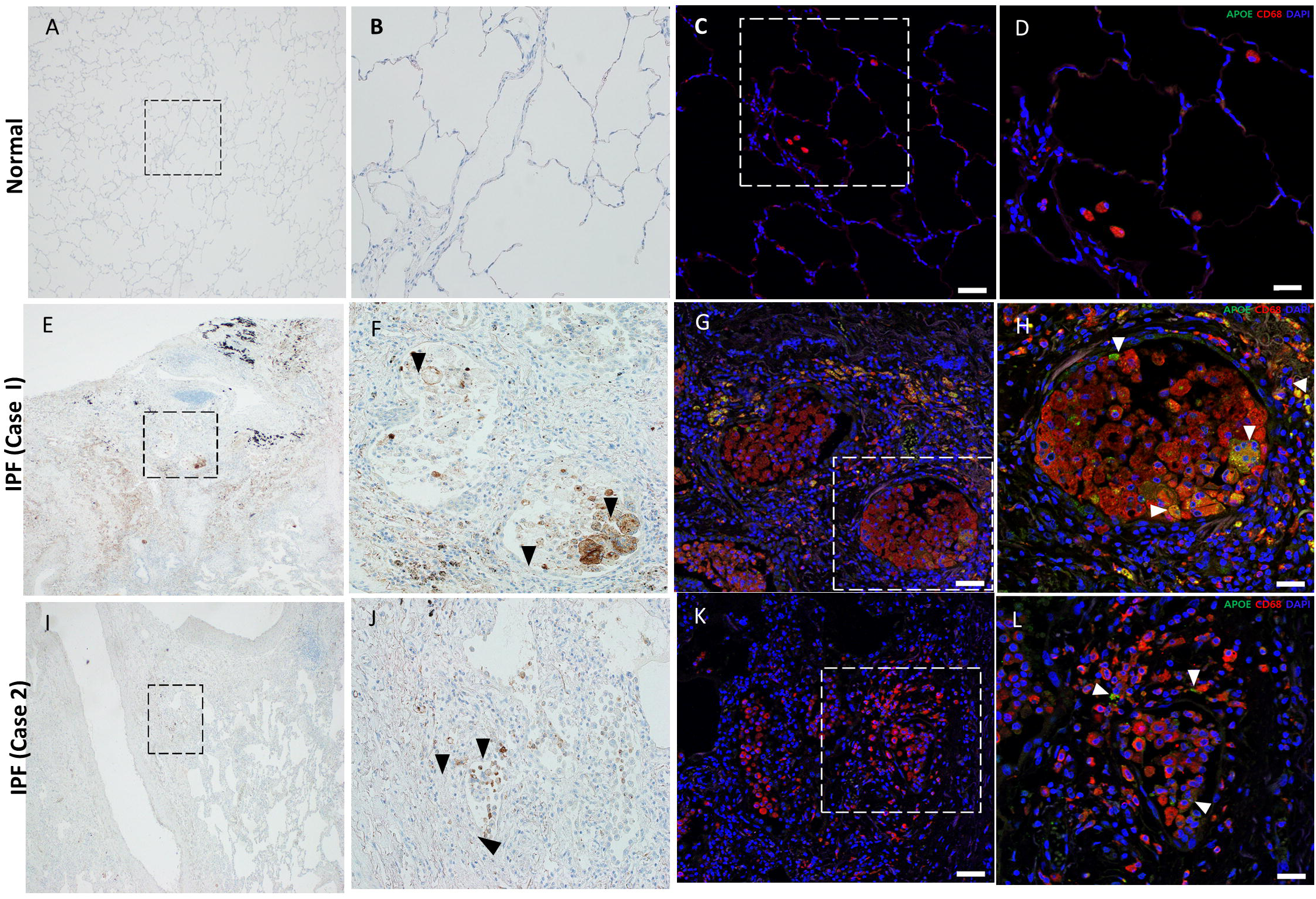
ApoE expression and co-expression with macrophage in normal and IPF lung tissues. (A, B) In the normal lung tissue, ApoE immunostaining revealed the absence of cytoplasmic ApoE expression. ApoE was not detected in the cellular cytoplasm after immunostaining. (C, D) Within the normal lung tissue, bronchioles showed exclusive expression of macrophages (shown in red) with no detectable cytoplasmic ApoE expression. (E, F, I, J) In contrast, in the IPF lung tissue of the patient, ApoE immunostaining exhibited robust and active ApoE expression, observed in the bronchioles or alveolar sacs (shown as black spots) and within the interstitium (black arrows). (G, H, K, L) Immunofluorescence staining was performed to elucidate the functional role of ApoE in the pathway from bronchioles to lung tissue and explore the intricate interplay of cells and proteins in the gas exchange region, as well as its involvement in inflammatory and immune responses with macrophages. The analysis revealed strong ApoE expression (shown in green) in the cytoplasm surrounding the nucleus (blue), colocalizing with the presence of macrophages (shown in red). Cell nuclei were counterstained with DAPI (blue); scale bar: 20 μm.

### ApoE validation in human IPF scRNA sequencing

Apo E expression was validated through transcriptomic analysis in the blood of 20 IPF patients and 5 normal subjects. Apo E is present in ResM, BMM, and all macrophages and was expressed in both IPF patients and normal patients. However, differences in Apo E expression were observed depending on the progression of IPF. BMM had low ApoE expression, but as the disease progressed, APOE expression increased, and ResM showed reciprocal changes.(Figure 8)

**Figure 8.**
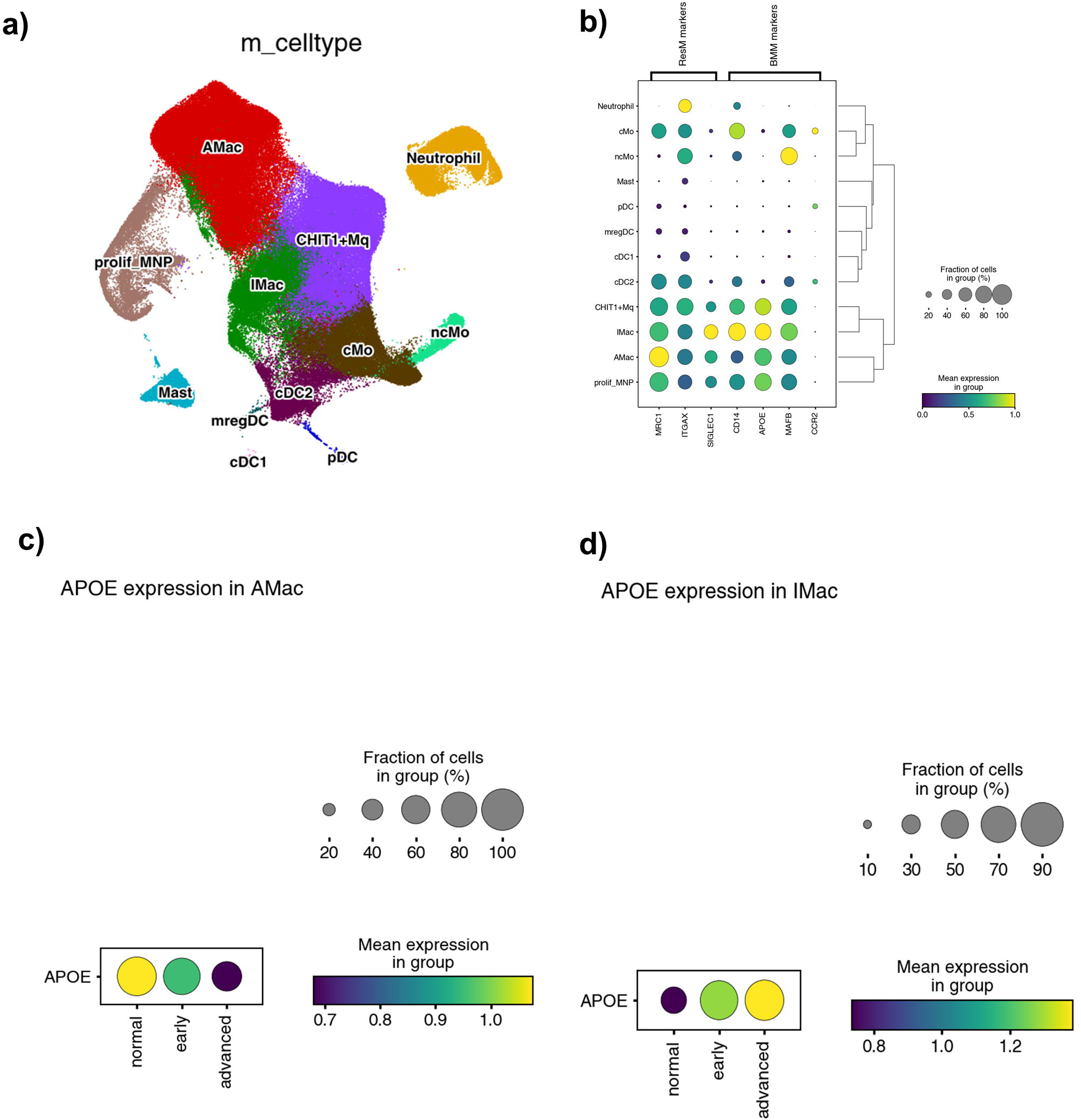
Apo E validation performed by scRNA sequencing of 20 IPF patients’ blood. A) Cell-type annotations projected into UMAP space B) UMAP plot of reclassified macrophage subclusters using ResM and BMM markers C) ApoE expression in alveolar macrophage (ResM) D) ApoE expression in interstitial macrophage (BMM)

## Discussion

In this study, we used a micellised nanoparticle type of prodrug BMP-7 targeting AM, which are primary regulators of the alveolar immune response. The antifibrotic effect of prodrug BMP-7 was demonstrated in a BLM model. We then used scRNA-seq of BALF to characterise macrophage heterogeneity during fibrosis. This transcriptome analysis enabled the subclustering of macrophages and revealed a fibrosis-associated subgroup with a distinctive gene expression profile, particularly bone marrow-derived ApoE+ AM. We validated these results through immunostaining in a BLM-induced mouse model with prodrug BMP-7 inhalation. Furthermore, we demonstrated the upregulation of ApoE and co-localised macrophages in human IPF lungs and validate Apo E expression in human IPF blood sample using scRNA seq.

Recent studies have reported that two ontological populations of AM coexist during pulmonary fibrosis: ResM and BMM, and BMM are known to have distinct functions during fibrogenesis [6, 9, 28]. These transcriptionally heterogeneous populations have distinct roles in pathogenesis, which have been researched through scRNA-seq. ApoE is one of the DEGs between ResM and BMM [10, 29].

ApoE is highly expressed in multiple subtypes of lung cells, including fibroblasts, AT2 cells, B cells and AM, accumulating during the injury response [10]. In a BLM-induced mouse model, but not ResM, primarily express ApoE and play a crucial role in the resolution of fibrosis by directly binding to collagen I, thereby mediating phagocytosis [30]. Additionally, upregulated ApoE macrophages promote muscle repair in a CXCR1-deficient mouse model by enhancing phagocytosis and restoring muscle regeneration [31].

When comparing the expression levels of ApoE among macrophage subpopulations, the expression in ResM was lower than in BMM; however, both subpopulations showed a similar pattern of increase in G3 and decrease in G4 compared to the control groups (G1, G2). This suggests that BLM-induced fibrosis was reduced by prodrug BMP-7 through the downregulation of ApoE expression in macrophages. Although ApoE has been reported to be expressed primarily in monocyte-derived macrophages, in this experiment, ApoE was expressed in both BMM and ResM.

Previous studies have indicated that the complement system plays a significant role in the process of fibrosis [32, 33]. Addis-Lieser et al. reported that C3 knockout mice demonstrate reduced BLM-induced lung injury compared to wild-type mice. Additionally, C5 has been suggested to play a profibrotic role. In this experiment, the C1qb gene increased in G3 and decreased in G4, which aligns with these findings and is supported by the GSEA results.

Moreover, previous studies indicate that fibrosis increases in the hearts of Mt1/2 knockout mice [34]. The decrease in Mt1/2 expression in G3, where fibrosis was induced, and the subsequent increase in G4 are consistent with previous research. Mt1 has various immunomodulatory effects, such as modulating several signal transduction pathways, including the phosphorylation of STAT signalling pathways and NF-kB signalling [35]. However, the role of Mt1/2 during fibrosis has been rarely reported. It is known to be associated with organ remodelling, with published data showing attenuated myocardial remodelling in Mt-overexpressing mice [36].

In the GSEA for BMM G4 vs. G3, the hallmark of the TGF-β pathway, was detected as the sixth lowest pathway with treatment of prodrug-BMP-7, indicating its potential relevance to inhibit the TGF-β pathway and attenuate pulmonary fibrosis. Other scRNA-seq studies have identified macrophages expressing upregulated profibrotic factors interacting with specific cytokines and chemokines related to fibroblast attractants [37]. A previous study has reported that TGF-β/Smad3 signalling regulates the transition of BMM into myofibroblasts during renal tissue fibrosis [38].

Therefore, while downregulation of ApoE in AM does not directly involve TGF-β signalling, it may affect fibrosis through interactions with fibroblasts or myofibroblasts, subsequently downregulating TGF-β signaling related to fibrosis.

The Kaminski group’s scRNA data compared APOE expression across the entire myeloid lineage according to IPF, normal, and COPD, and reported that Apo E expression was highest in IPF, showing similar findings to our study[39].

Although differences in the subpopulation proportions of lymphocytes were observed between experimental groups, the DEG analysis showed no significant differences at the molecular level, leading to their exclusion from further analysis.

A limitation of this study is human macrophages are difficult to obtain, and because human and mouse macrophages differ, research results may not always translate directly to clinical settings.

In conclusion, our finding suggests that inhibition of the TGF-β/smad2/3 pathway through AMs, especially ApoE macrophage, could be a therapeutic target for treatment of pulmonary fibrosis. And Prodrug BMP-7, which modulate ApoE alveolar macrophage could be suitable for this process. Further research is required to determine whether prodrug BMP-7 suppresses the inflammation or fibrosis stages, and additional functional study with ApoE knock out mice are needed.

## Data Availability

All data produced in the present study are available upon reasonable request to the authors

## Acknowledgments

This work was supported by grants from the National Research Foundation of Korea (NRF-2021R1A2C3003496, NRF-2022R1A2C3004609, NFR-2022M3A9G8016257) funded by the Korean government (MSIP).

## Conflict of Interest

NHK, HSK, and JIY are the founders of MET Life Sciences Co., Ltd., and shareholders. The remaining authors declare that the research was conducted in the absence of any commercial or financial relationships that could be construed as a potential conflict of interest.

## Abbreviation

IPF: idiopathic pulmonary fibrosis
scRNA-seq: single-cell RNA sequencing
AEC: alveolar epithelial cell
EMT: epithelial mesenchymal transition
TGF-β: transforming growth factor β
AM: alveolar macrophages
IM: interstitial macrophages
ApoE: apolipoprotein E
PTD: protein transduction domain
ICG: indocyanine green
BMP: bone morphogenic proteins
rhBMP-7: recombinant BMP-7
prodrug BMP-7: micellised protein transduction domain bone morphogenic protein-7
BLM: bleomycin
BALF: bronchoalveolar lavage fluid
BMM: bone marrow-derived macrophages
ResM: resident macrophages

## Notes

### Competing Interest Statement

The authors have declared no competing interest.

### Author Declarations

Institutional Review Board of Severance Hospital gave ethical approval for this work.

