## supplementary documents for "Prodrug BMP-7 attenuates pulmonary fibrosis through downregulation of bone marrow derived ApoE+ alveolar macrophage"

**Isolation of bronchoalveolar lavage cells and cell counts**

At 21 days after BLM/PBS instillation, all mice were humanely euthanized by lethal overdose of ketamine and xylazine. BALF was collected through a tracheal cannula using two 1 mL aliquots of sterile saline. Samples were then centrifuged for 10 minutes at 4 °C and 1,500~5,000 rpm. The resulting cell pellet was resuspended in 100 μL PBS, and analyzed by quantitative and qualitative cell counting. According to the manufacturer’s protocol, cells were counted using a hemocytometer (Marienfield, Germany). Slide chambers were prepared by inserting slides into frames with the poly-L-lysine coating up and clamping with clips on either side. A 90 ul aliquot of each sample was inserted into a cytospin with slides facing outward, filled with 90 μL sample, and centrifuged at 600 rpm for 6 minutes. The slides were then retrieved, dried, and stained by immersing in Diff Quick (Sysmex Corporation) The slides were immersed in three Diff Quickfluids (Fixative, Solution I, and Solution II) for 5 seconds and rinsed with purified water. The protein content of the BAL supernatant was measured using the Coomassie Brilliant Blue G-250 technique (Quick StartTMBradford Protein Assay). 25 ul of each sample and 200 μl of working reagent were pipetted into a microplate well and mixed thoroughly on a plate shaker for 30 s. After incubation for 30 min at 37◦C, the plate was cooled and the absorbance read at 562 nm in a spectrophotometer.

**Lung tissue harvest and histologic examination**

The right lung was isolated and stored at −80◦C prior to protein extraction, after flushing the pulmonary vasculature with saline under low pressure. The left lung was inflated via the tracheotomy with low-melting point agarose (4%) in PBS at 25 cm H_2_O pressure and until the pleural margins became sharp. Tissues were then excised, fixed overnight in 10 % formaldehyde in PBS, embedded in paraffin, and sectioned at 5 μm. The sections were then stained with hematoxylin/eosin and Masson's trichrome, and scored under a light microscope by two qualified investigators blinded to samples. Fibrotic changes in lung sections were graded semi-quantitatively according to the scale defined by modified Ashcroft score.[27] All of the sections of the lung parenchyma were assessed using a score of 0- 8. In every field, fibrosis was recorded as predominant when it occupied more than half of the area. The final score was expressed as the mean of the individual scores observed on all fields.

**Western blotting**

The Frozen lungs were mechanically homogenized in 600 μL with homogenization buffer (PRO-PREPTM Extraction Solution), cell lysed induced by incubation for 20-30 minutes on ice or at -20 °C, and centrifuged at 13,000 *×g* for 30 minutes at 4 °C. Equal amounts of protein were separated by SDS-PAGE, and transferred to nitrocellulose membranes before immunoblotting with primary antibodies. Membranes were then incubated with anti-rabbit or anti-mouse IgG conjugated to horseradish peroxidase conjugated, visualized using Super-Signal West Pico Chemiluminescence Detection Kit (Pierce), The band images were quantified using in Alpha Ease FC version 4.1.0 (Innotech). The antibodies used in the present study included α-SMA (abcam #ab7817, 1: 2,000), Collagen I, (abcam #ab34710, 1: 2,000), Fibronectin (abcam #ab2413, 1: 2,000), β-actin (santa cruz #sc-47778, 1: 1,000), p-smad2/3 (cell signaling #8828, 1:500), Smad2/3 (cell signaling #3102, 1:500).

**Single cell RNA-seq analysis of BAL fluid cells**

BAL fluid single cells were collected and pooled for each group. In total 40,000 of cells per group were loaded onto microwell cartridges of the BD Rhapsody Express system (BD). Single cell whole transcriptome analysis libraries were prepared using BD Rhapsody WTA Reagent kit (BD,633802) according to the manufacturer’s instruction. The final index PCR libraries were sequenced on the Illumina HiSeq using High Output Kit v2.5 (150 cycles, Illumina) for 2 x 75 bp paired-end reads with 8 bp single index.

The FASTQ-format sequencing raw data was processed with BD Rhapsody WTA Analysis pipeline (version 1.0, Revision 6) on SevenBridges Genomics online platform (SevenBridges) and expression matrix was used for further data analysis. Data normalization, dimensionality reduction and visualization were performed using Seurat package (version 4.3.0) unless otherwise specified.

To ensure data quality, cells were filtered based on the criteria: having a number of genes per cell (nFeature_RNA) between 500 and 6000, and a percentage of mitochondrial genes (percent.mito) less than 25. Additionally, genes were filtered to include only those present in a minimum of 3 cells. After filtering, the matrices were normalized using the NormalizeData function with the LogNormalize method and a scale factor of 10,000. Variable genes were identified using the FindVariableFeatures function, selecting the top 2000 genes with the variance stabilizing transformation (VST) method, while also excluding genes related to the cell cycle (GO:0007049). Data integration was achieved through FindIntegrationAnchors and IntegrateData functions with default options. The integrated data was further processed using the ScaleData and RunPCA functions. Statistically significant principal components were identified using the JackStraw method, which were then used for UMAP non-linear dimensional reduction.

For unsupervised hierarchical clustering analysis, the FindClusters function in Seurat package was applied. Various resolutions between 0.1 and 0.9 were tested, and the final resolution was selected based on the most stable and relevant outcome, utilizing the clustree R package and considering prior knowledge. Cellular identity for each cluster was determined by finding cluster-specific marker genes using the FindAllMarkers function. Marker genes were considered with a minimum fraction of cells expressing the gene over 25% (min.pct=0.25), and comparisons were made against known cell type-specific genes from PanglaoDB (https://panglaodb.se).

Differentially expressed genes were identified using the FindMarkers function of the Seurat package with logfc.threshold = 0. Gene set enrichment analysis was performed using the Molecular Signatures Database (MSigDB) with the fgsea R package (Korotkevich, G., Sukhov, V. & Sergushichev, A. Fast gene-set enrichment analysis. Preprint at BioRxiv https://doi.org/10.1101/060012 (2019)). Differentially expressed genes between different groups were identified using the FindMarkers function in Seurat with min.pct = 0.25 and logfc.threshold = 0, pre-ranked according to the average log2 fold change values and used for enriched gene set identification using the gfsea function with minSize = 15, maxSize = 500, and nperm = 1000 options.

**Immunofluorescence & Immunohistochemistry staining**

Unstained mouse lung tissue slides embedded in formalin-fixed 5 μm paraffin blocks were deparaffinized and washed three times with PBS. Subsequently, the slides were incubated in 1% citrate buffer (pH 8.0) at 95-98°C for 45 minutes to facilitate antigen retrieval. After two additional washes with PBS, permeabilization was performed using 1% SDS for 10 minutes.

Following permeabilization, the primary antibodies, APOE recombinant rabbit monoclonal (ThermoFisher Scientific, catalog number 701241), and the macrophage marker, CD68 (SANTA CRUZ BIOTHERAPY, INC, catalog number sc-20060), were diluted in DAKO antibody dilution buffer (1:1,000) and incubated overnight at 4°C. Goat anti-rabbit IgG secondary antibody, Alexa FluorTM 488, or goat anti-mouse IgG secondary antibody, Alexa FluorTM 568 (Invitrogen), was diluted 1:1,000 and incubated for 1 hour at room temperature. The secondary antibodies were then washed with PBS, and the nuclei were stained with 1 μg/ml DAPI for 20 minutes.

Fluorescence images were captured using a Zeiss LSM700 confocal microscope (Carl Zeiss, Berlin, Germany). Morphological analysis of the acquired confocal images was performed using Metamorph microscopy analysis software (version 7.1, Molecular Devices, Sunnyvale, CA, USA). Twenty-four-bit confocal images were used for image quantification in each condition.
