## supplementary figure for "Prodrug BMP-7 attenuates pulmonary fibrosis through downregulation of bone marrow derived ApoE+ alveolar macrophage"

### Slide 1
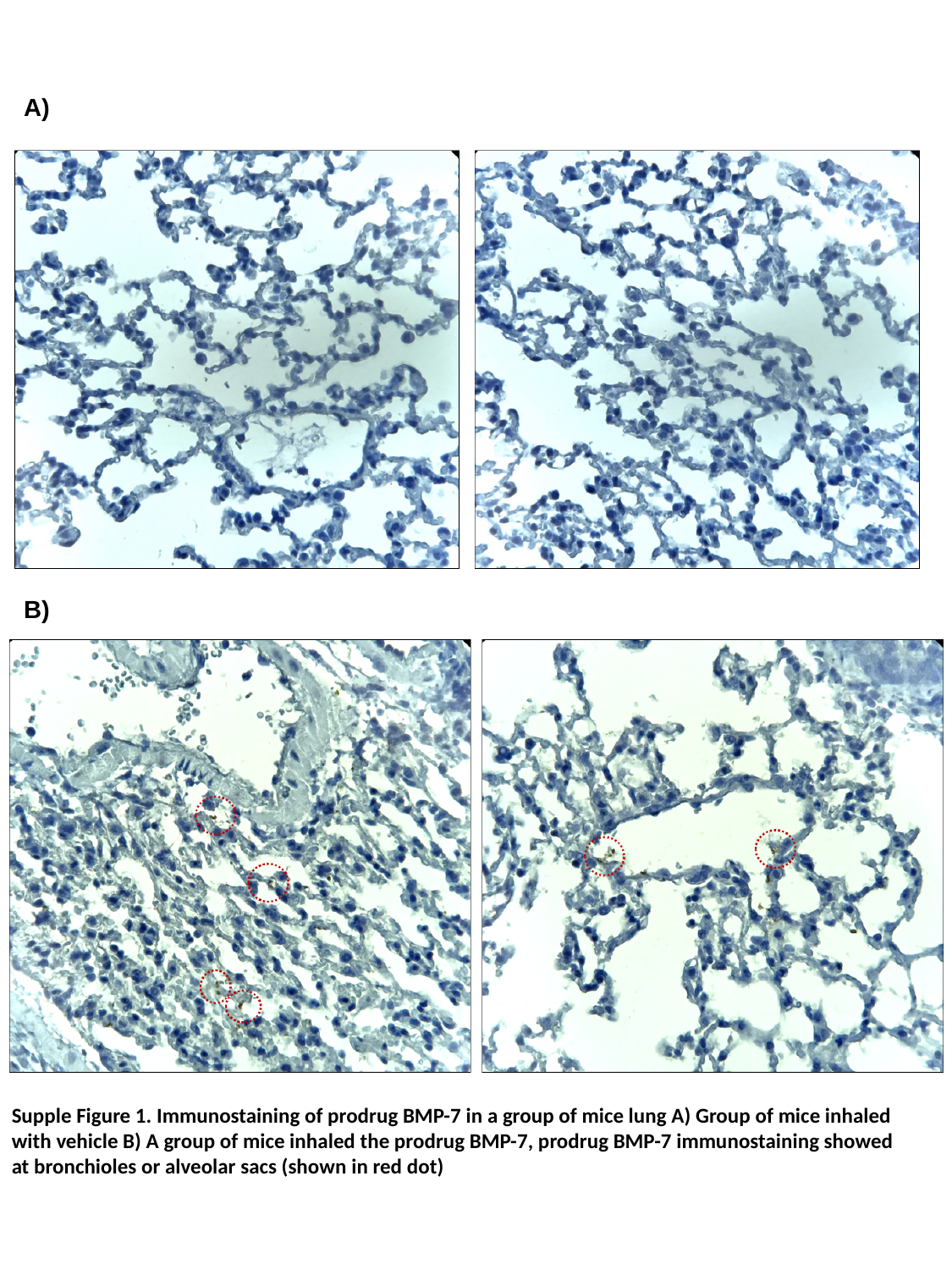

A)
B)
Supple Figure 1. Immunostaining of prodrug BMP-7 in a group of mice lung A) Group of mice inhaled with vehicle B) A group of mice inhaled the prodrug BMP-7, prodrug BMP-7 immunostaining showed at bronchioles or alveolar sacs (shown in red dot)

### Slide 2
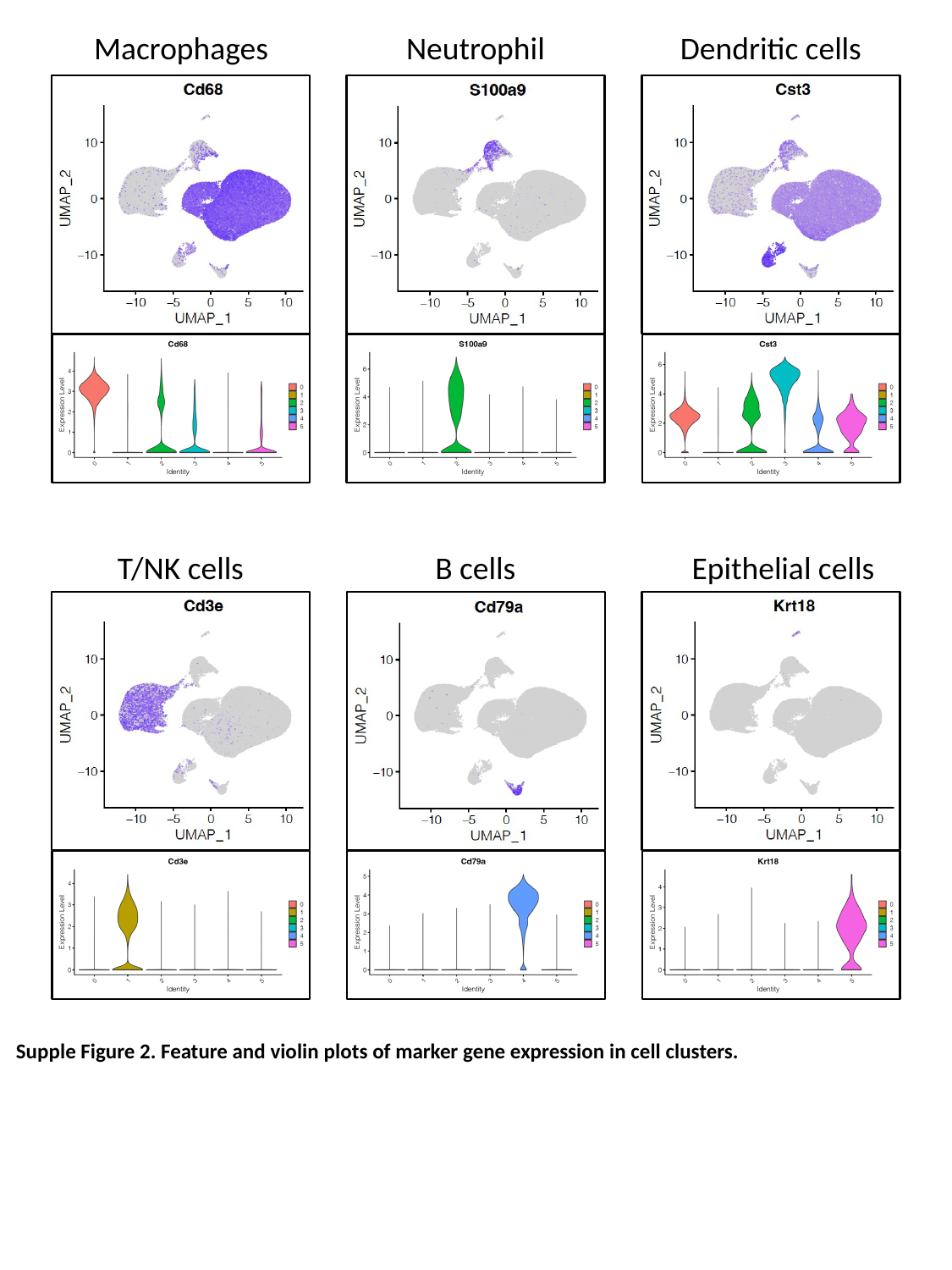

Macrophages
Neutrophil
Dendritic cells
B cells
Epithelial cells
T/NK cells
Supple Figure 2. Feature and violin plots of marker gene expression in cell clusters.

### Slide 3
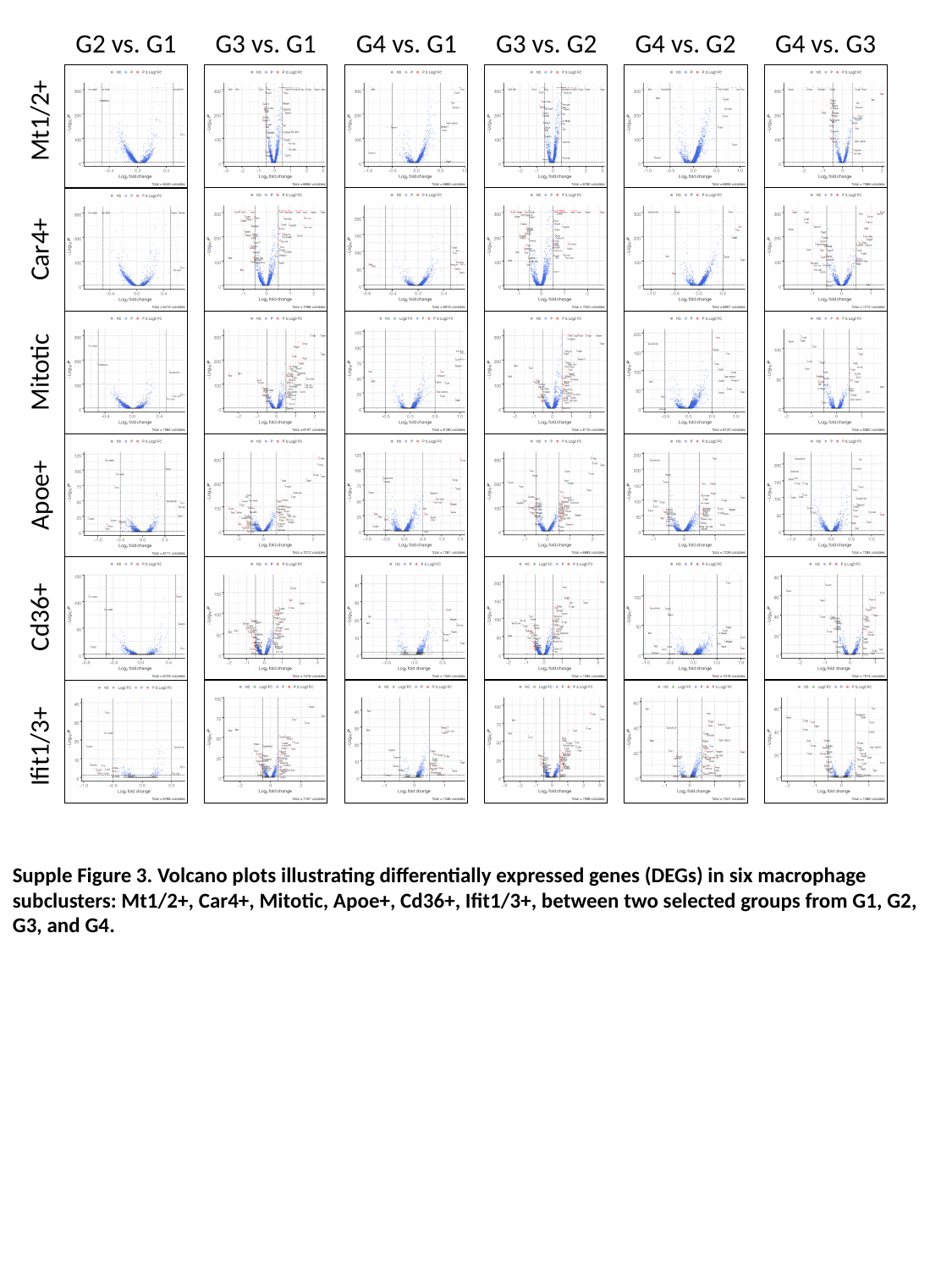

G2 vs. G1
G3 vs. G1
G4 vs. G1
G3 vs. G2
G4 vs. G2
G4 vs. G3
Mt1/2+
Car4+
Mitotic
Apoe+
Cd36+
Ifit1/3+
Supple Figure 3. Volcano plots illustrating differentially expressed genes (DEGs) in six macrophage subclusters: Mt1/2+, Car4+, Mitotic, Apoe+, Cd36+, Ifit1/3+, between two selected groups from G1, G2, G3, and G4.

### Slide 4
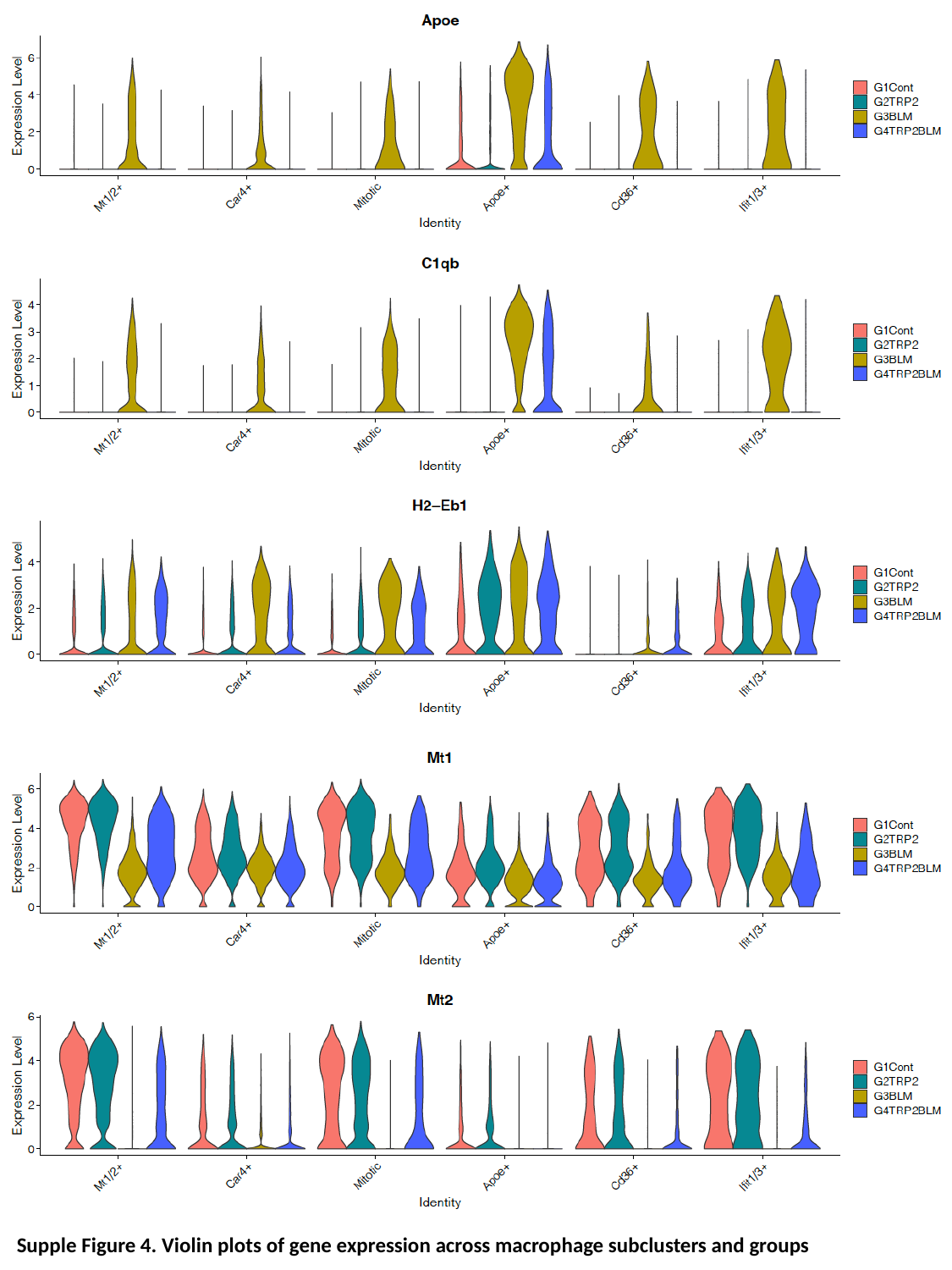

Supple Figure 4. Violin plots of gene expression across macrophage subclusters and groups

### Slide 5
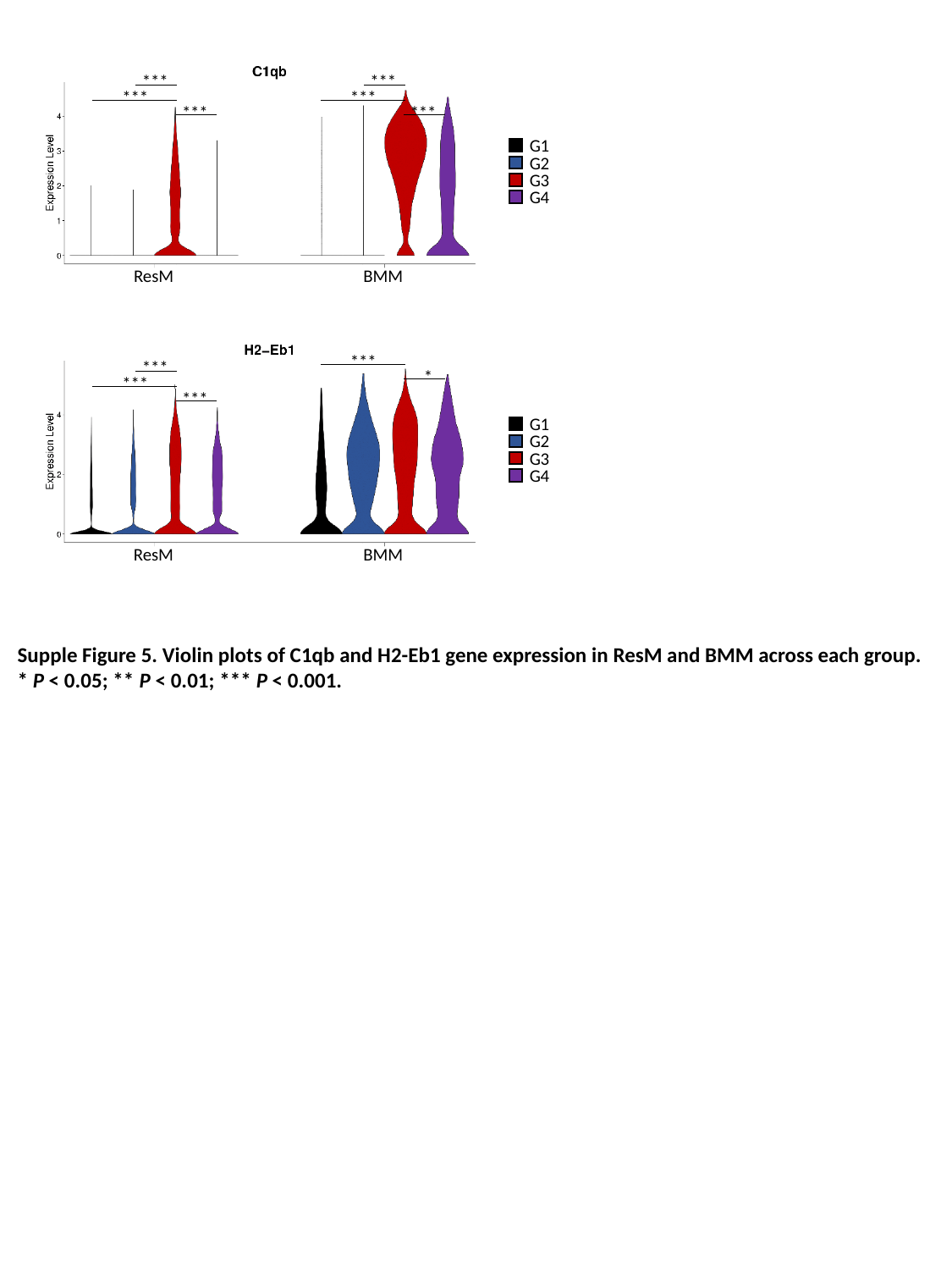

BMM
ResM
G1
G2
G3
G4
***
***
***
***
***
***
ResM
BMM
G1
G2
G3
G4
***
*
***
***
***
Supple Figure 5. Violin plots of C1qb and H2-Eb1 gene expression in ResM and BMM across each group.
* P < 0.05; ** P < 0.01; *** P < 0.001.

### Slide 6
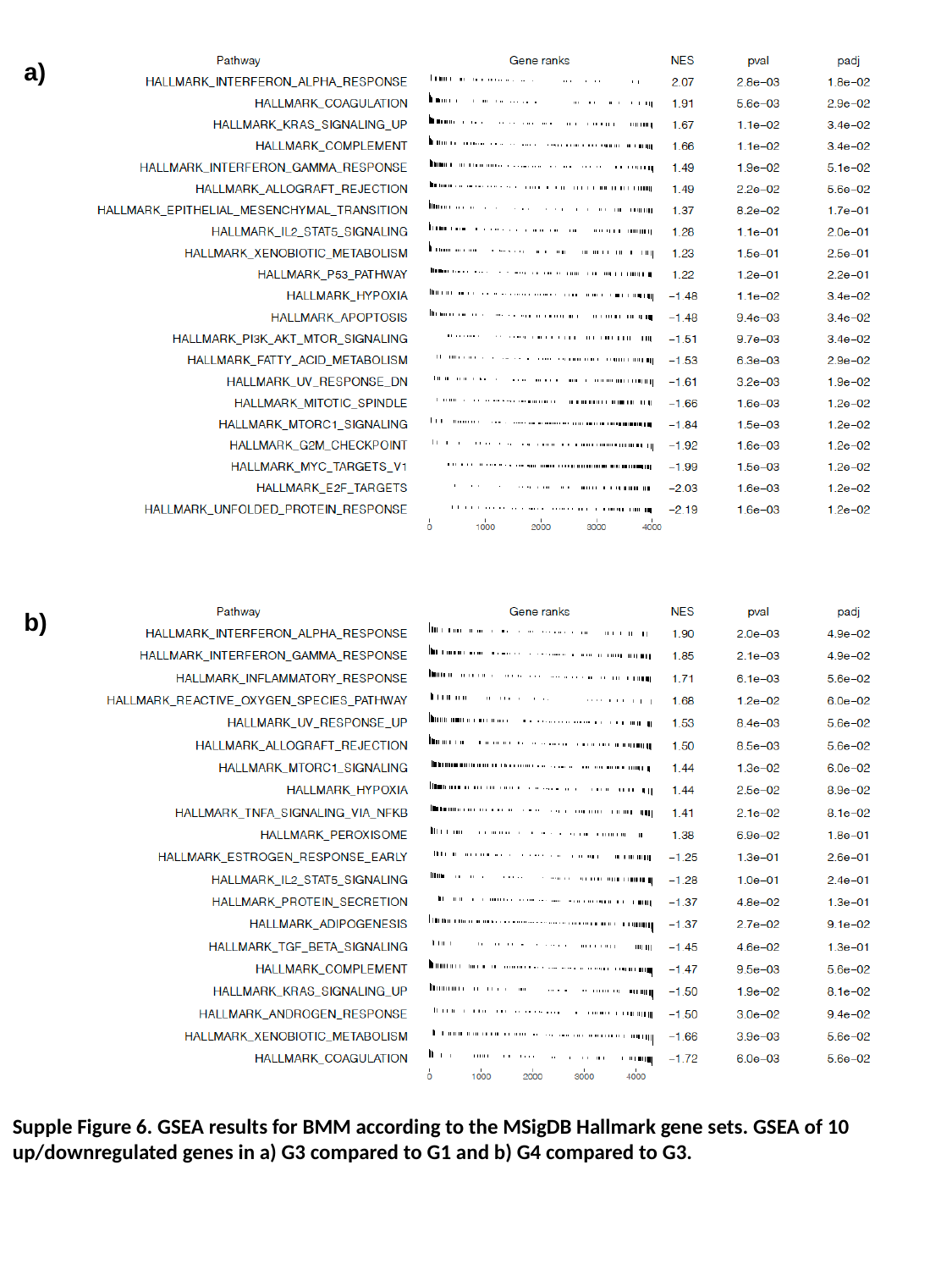

a)
b)
Supple Figure 6. GSEA results for BMM according to the MSigDB Hallmark gene sets. GSEA of 10 up/downregulated genes in a) G3 compared to G1 and b) G4 compared to G3.

### Slide 7
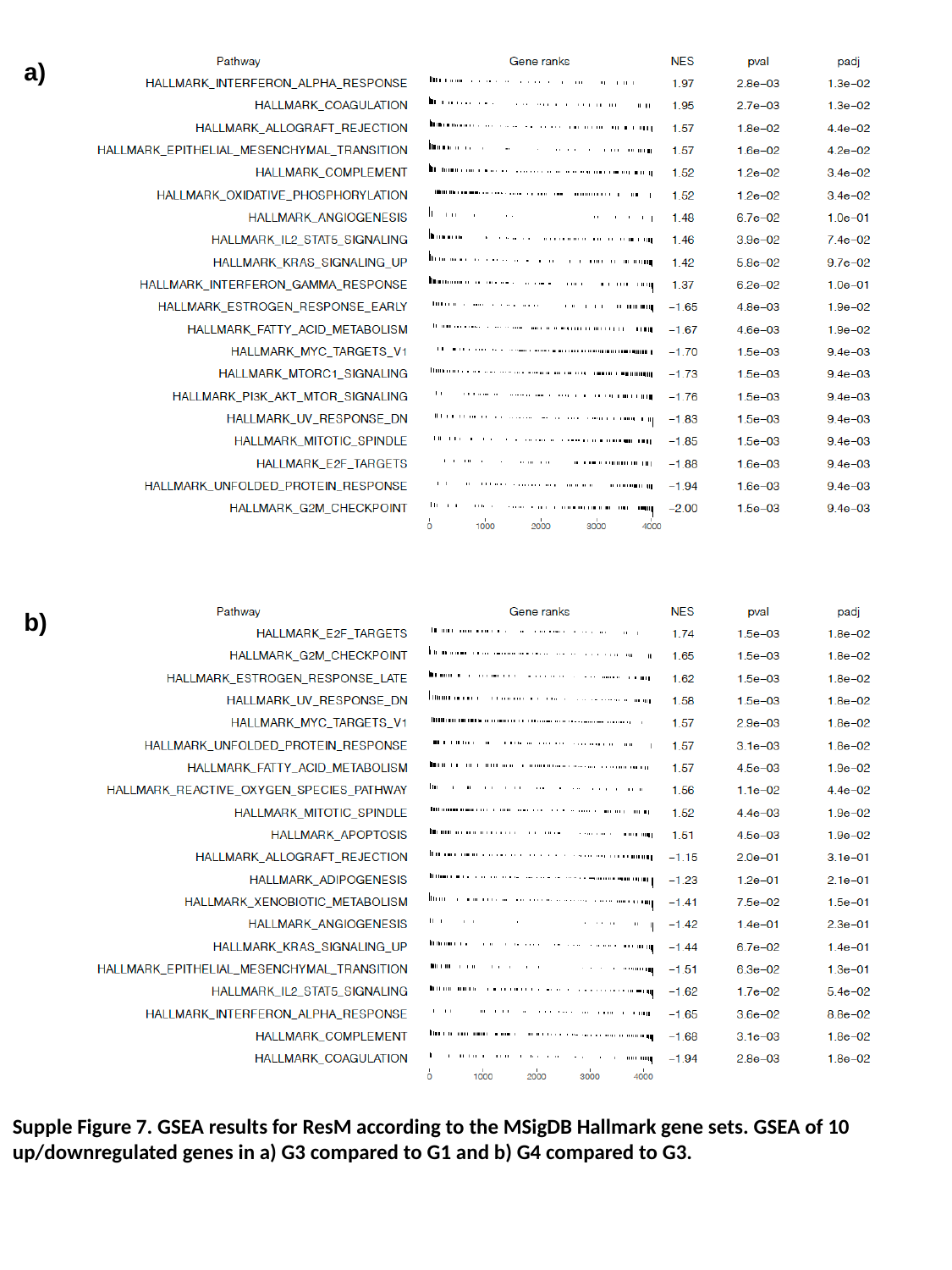

a)
b)
Supple Figure 7. GSEA results for ResM according to the MSigDB Hallmark gene sets. GSEA of 10 up/downregulated genes in a) G3 compared to G1 and b) G4 compared to G3.
